# On the Impact of Mass Screening for SARS-CoV-2 through Self-Testing in Greece

**DOI:** 10.1101/2023.02.15.23285963

**Authors:** Samuel Gilmour, Spyros Sapounas, Kimon Drakopoulos, Patrick Jaillet, Gkikas Magiorkinis, Nikolaos Trichakis

## Abstract

The emergence of COVID-19 stressed country health systems up to the point of triggering compulsory public health interventions to flatten the epidemic curve. Most of the interventions during the first year of the pandemic were non-pharmaceutical and aimed to reduce the contact rate of the people, which reduced the transmission rate of all respiratory pathogens, but had a large social and financial burden. SARS-CoV-2 specific interventions included screening, that is testing of asymptomatic people, which was largely facilitated by the availability of self-testing lateral flow antigen detection devices. The importance of self-testing interventions in controlling COVID-19 epidemic is not well-documented. We study as a paradigm-model the self-testing COVID-19 mass screening program that was implemented in Greece, involving large, susceptible populations taking tests routinely and pre-emptively so as to enable early detection of infections. Using a novel compartmental model we quantify the effectiveness of the program in curbing the COVID-19 pandemic. Conservative estimates indicate that the program reduced the reproductive number by 4%, hospital admissions by 25% and deaths by 20%, which translated into approximately 20,000 averted hospitalizations and 2,000 averted deaths between April-December 2021. Self-testing mass screening programs are efficient interventions with minimal social and financial burden, thus they are invaluable tools to be considered in pandemic preparedness.

---

The global emergence of the COVID-19 pandemic caught the entire world off guard with devastating effects. During the years 2020-21, widespread community transmission of the SARS-CoV-2 pathogen frequently resulted in unprecedented demand for healthcare resources that stretched entire national health systems beyond their capacity across the world. In response to this public health emergency, governments and local authorities adopted different intervention strategies in their attempts to curb COVID-19 spread, ranging from public advisories to face mask mandates, travel restrictions, and strict lockdowns.

In this work, we study the novel COVID-19 mass screening program that was implemented in Greece. Mass screening programs involve large, susceptible populations taking tests routinely and preemptively so as to enable early detection of infections. Our goal is to quantify the effectiveness of the Greek mass screening program in curbing the COVID-19 pandemic, particularly its impact on averting hospitalizations and saving lives. We also seek to obtain insights on best practices and lessons learned from the operational decisions that were made. The analysis could help inform policymakers who contemplate mitigating strategies, particularly whether to roll out screening programs, in times of future pandemics. The choice of what strategy to follow and which interventions to mount in order to combat the COVID-19 pandemic presented policymakers with perplexing dilemmas. The associated calculus often revolved around balancing the expected negative and positive consequences for each course of action. The former related to a wide range of potentially harmful implications, such as direct financial costs, disrupted education, adverse effects on mental health, among many others. The latter primarily related to the intervention’s effectiveness in ultimately relieving pressure on the health system and saving lives.

Predicting the consequences of an intervention during COVID-19 was, of course, in and of itself a very challenging problem. For some interventions that had been adopted in previous respiratory pandemics similar to COVID-19, extant documentation in the literature, albeit limited, provided guidance and invaluable experience. Analyses of novel interventions with no historical precedence prior to COVID-19 could therefore prove vital in informing how we respond to future crises.

Mass screening programs as mitigating interventions for infectious diseases, and particularly for acute viral infections, were implemented for the first time during the COVID-19 pandemic. In particular, technological and scientific advancements in diagnostics, coupled with robust manufacturing and supply chain capabilities, enabled few countries, including Greece, to procure and distribute self-testing kits to the public at a massive scale.

The Greek mass screening or self-testing program was initiated in April 2021 and was rolled out in different phases. The program was targeted at several population groups in Greece: students, teaching staff, civil servants, and private sector employees. For the most part, each individual within the targeted groups was required by law to take two self tests per week, regardless of symptoms of disease, and report the results at a centralized online platform. The entire population was also encouraged to take tests, and indeed free kits were occasionally distributed to everyone, particularly preceding or following holidays that typically involved large gatherings of people. The selection of self-testing kits was based on the rate of sensitivity (above above 85% for samples testing positive with a qPCR up to the 33rd reaction cycle and with a specificity of over 99%) with a specificity of over 99% which was supported through peer reviewed process or evaluation available at FIND’s^1^ test directory. Importantly, these tests were also safe, inexpensive, acceptable by the target population and simple to take at home or anywhere, providing rapid results. More implementation details of the program are provided as Supporting Information.

When it comes to ascertaining the negative and positive consequences of mass screening programs, the former are typically much easier to quantify a priori. Indeed, the cons are primarily limited to direct financial costs, such as procurement and distribution costs, that can be reasonably estimated prior to implementation. The expected pros, however, are more challenging to assess, as they stem from early detection and isolation of infected individuals, including asymptomatic ones, which in turn could help limit disease transmission. Therefore, our analysis of the Greek program focuses solely on quantifying its benefits.

Our study attempts to quantify the impact that the program had on curbing the COVID-19 pandemic in Greece between April 4, 2021, the date that program was launched, and December 15, 2021, the date we have data until. During that period, a total of 60 million self-testing kits were distributed. At its peak during the study period, an estimated percentage as large as 20% of the population took two self tests over a week as part of the program.

## 1. Results

To analyze the impact of the Greek self-testing program, we developed a novel compartmental model that tracks the evolution of the COVID-19 pandemic in Greece. The model is reminiscent of an *SIR* model, but with important modifications that enable us to model salient dynamics, such as the testing and vaccination operations in Greece. At a high level, the compartments model susceptible individuals, who might become infected and contagious after being exposed to the virus. Infected individuals can be asymptomatic or symptomatic before either recovering with immunity or dying. Infected individuals might also be identified through testing, in which case they are isolated to avert disease transmission. To capture the effects of the self-testing operations, which were allocated in different proportions to age groups 0 to 18 years, 19 to 64 years, and 65+ years, we consider three sets of compartments that correspond to the said age groups. Depending on which age group they model, we index compartments by *a*, which takes values in 𝒢 = ‘0-18’,’19-64’,’65+’. Similarly, we also consider two sets of compartments based on vaccination status, indexed by *v* ∈ {0, 1} to indicate vaccination. Figure 1 details, for a single age group *a* ∈ 𝒢, the model compartments and the possible transitions among them.

**Fig. 1.**
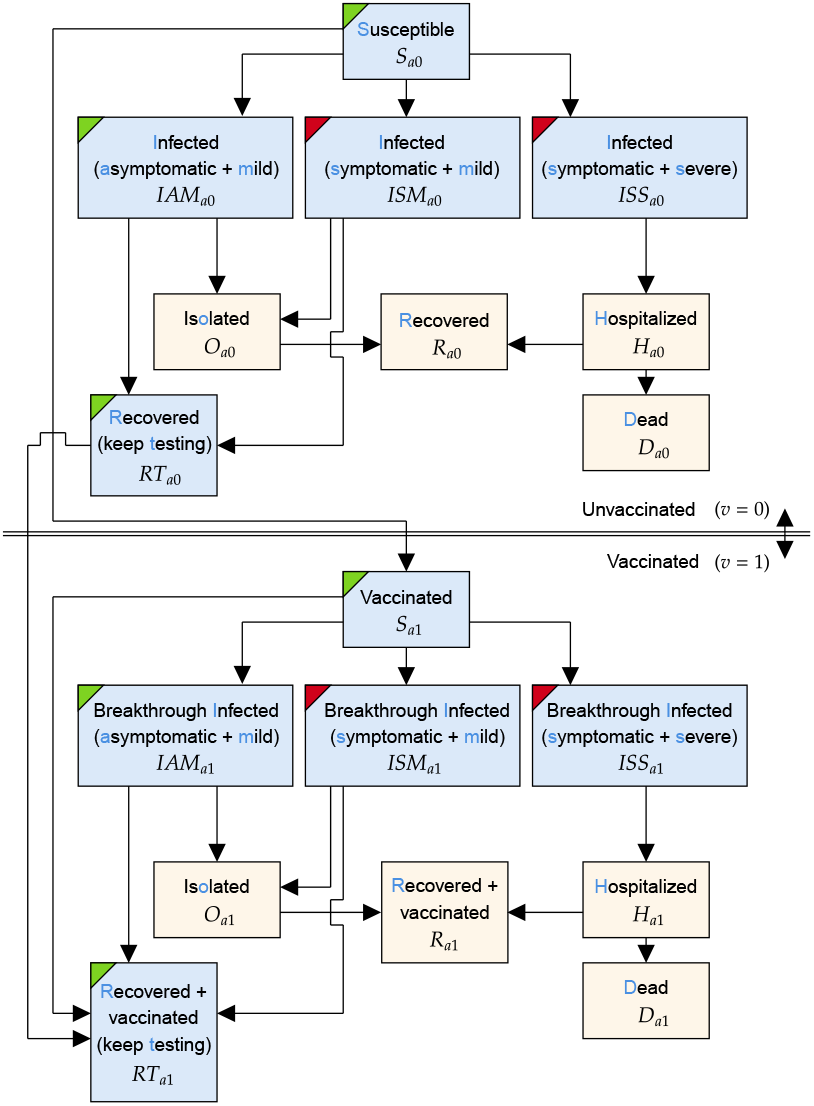
The structure of the model within a particular age group *a* ∈ 𝒢. For some vaccination group *v* ∈ *{*0, 1*}*, the states within this age group are indexed by subscript *{av}*. Compartments colored with a blue background are compartments that are subject to testing. Compartments with green and red corners contain asymptomatic and symptomatic individuals, respectively.

The model was fitted to highly detailed historical data that we obtained from the Greek National Public Health Organization. Further details on the model, the data, and the methods used are provided in Section 3.

### A. Overall Impact of the Program

A high-level description of how we evaluate the overall impact of the self-testing program is as follows. We use the fitted model, but modify the number of distributed self tests and set it to zero. After this modification, and keeping other parameters constant and equal to their fitted values, we simulate the model to obtain trajectories that the pandemic in Greece would have followed, had the self-testing program not been implemented. A rigorous presentation of the approach can be found in Section E.

We first evaluate the impact of the self-testing program on reducing the effective reproduction number (*R*_*t*_) of the pandemic. To conduct this analysis, we utilize the methodology of calculating *R*_*t*_ proposed in (1); see Section E for details. Table 1 presents the 80% confidence interval for the average reduction of *R*_*t*_ and the maximum weekly reduction over the period of study. The analysis suggests the program reduced the virus’ transmissibility by 4.7% on average, while the largest weekly reduction was 24%, approximately.

**Table 1.** Estimates for the percentage reduction in *R*_*t*_, and absolute reduction in deaths and hospitalizations due to the self-testing program in Greece (values in parentheses provide 80% confidence intervals).

| Metric | Estimate |
| --- | --- |
| Percentage reduction in $R_t$ | |
| Mean | 4.72 (3.93 – 5.37) |
| Maximum | 24 (17.3 – 25.6) |
| Reduction in deaths |  |
| Using direct method | 4888 (3698 – 6808) |
| Using indirect method | 3434 (2350 – 4387) |
| Reduction in hospitalizations |  |
| Using direct method | 40655 (33625 – 50699) |
| Using indirect method | 28379 (21763 – 34425) |

Next, we evaluate the impact of the program on deaths and hospitalizations. To further enhance precision, we employ two methods to conduct this analysis. The first is a ‘direct’ application of the methodology we described above, in which we perturb the fitted model by setting the number of self-tests to 0 to provide an estimate. The second is an ‘indirect’ application of the methodology we described above, in which we use only local perturbations of the fitted model (± 1% changes in the number of self-tests) and arguments from convex analysis to provide an estimate. Details are again deferred for Section E.

Table 1 presents the 80% confidence intervals on the number of deaths and hospitalizations that were averted by the self-testing program, as obtained by the direct and indirect methods of estimation. For reference, the total number of deaths observed in the historical data over the self-testing period was 10336, and the total number of hospitalizations was 76299.

Our most conservative estimates on the effect of the implemented self-testing program suggest mortality reduction of at least 20%, which corresponds to 2, 000 deaths, approximately. Furthermore, the program yielded a reduction in hospital admissions of at least 25%, which corresponds to 20, 000 hospitalizations, approximately. These additional hospitalizations would have stressed the Greek national healthcare system even further. It is important to note that our model does not consider the potential effect of this additional healthcare stress on deteriorating hospitalization outcomes and additional deaths, thus our mortality estimate is conservative with respect to the dynamic status and responsiveness of the healthcare system.

Previous studies have shown that non-pharmaceutical interventions, apart from lockdowns, during the first wave reduced transmissibility by around 2-3%, approximately (2). These included banning of public events and school closure. In comparison, the self-testing program in Greece appears to have been at least equally effective in reducing transmissibility. It was also fairly effective in reducing averting deaths while reducing stress to the national healthcare system. Given that self-testing yields a cost that can be met with less than C1 per test, and that the aforementioned interventions, such as school closures, are associated with a much higher societal and financial burden, the self-testing screening program had a high-effectiveness/low-cost profile.

### B. Impact of Operational Decisions

We next focus on some key operational decisions that are involved when designing a mass screening program:

i. Scale: how many tests should be ordered?
ii. Target: which subpopulations should be targeted?
iii. Accuracy: how important is the clinical accuracy of the self-tests used?

Below, we discuss the effect of each of these dimensions separately.

#### Scale: Impact of the number of tests

During the timeframe of our study on average 2.1% of the population was tested daily, with over 60 million kits distributed in total. In Figure 2, we provide estimates on the percent change in deaths and hospitalizations had the program been scaled up or down by some factor, compared to the actual implementation in Greece. If the tests administered had been 20% fewer, for example, total deaths would have increased by 5%, and total hospitalizations would have increased by 8%, approximately, during the period of our study. The technical details of how these estimates were produced can be found in Section D.

**Fig. 2.**
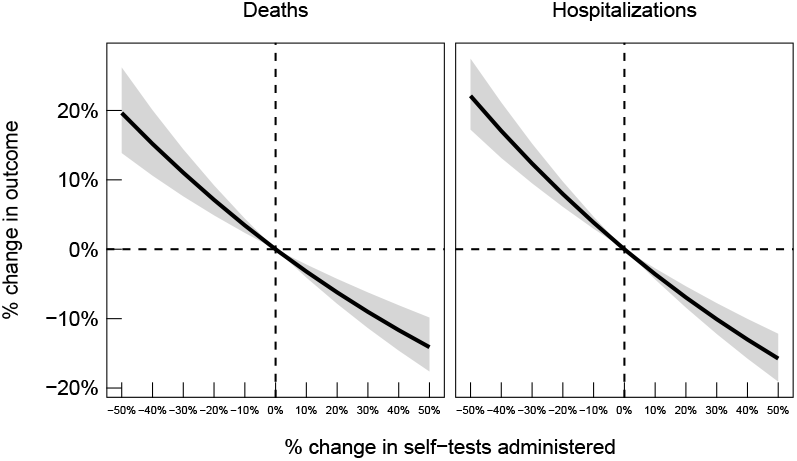
80% confidence intervals for the percent change in deaths (left panel) and hospitalizations (right panel) as a function of the percent change in the self-tests administered, relative to what is observed in the data.

Notably, the scale of the program, as measured by the number of tests, appears to yield diminishing reductions of deaths and hospitalizations. Therefore, on the one hand, more deaths and hospitalizations would have been averted, had the program been scaled up and more tests been administered. On the other hand, had the program been scaled down and fewer tests been administered, disproportionally more deaths and more hospitalizations would have occured, as the slopes of the curves appear to be steeper to the left of the implementation point, i.e., 0% change on the x-axis, than to the right.

#### Target: Impact of targeting subpopulations

In April 2021, when another wave of the pandemic was putting high pressure to the Greek National Health System, the authorities decided to launch the provision of two self-testing kits per week, for all students as well as the schools’ teaching and other staff. Later, in May 2021, private sector employees and civil servants who were not vaccinated or have not had COVID-19 were required by law to carry out two weekly tests. On average, 56.2% of the self tests was allocated to the 0 − 18 age group, 43.3% was allocated to the 19−64 age group and 0.5% was allocated to the 65+ age group.

In our analysis, we modified the fitted model to consider all alternative distributions of self tests amongst age groups — in other words, every possible three-way allocation of the total self tests across the age groups in our model. The best distribution was different for deaths and hospitalizations, though in both cases the percentage reduction over the observed distribution was small. For deaths, 30% of self-tests allocated to the 0 — 18 age group and 70% to the 19 — 64 age group resulted in a 2.23% reduction (80% CI: -3.85 – 6.84 %). For hospitalizations, 40% of self-tests allocated to the 0 – 18 age group and 60% to the 19 – 64 age group resulted in a 1.16% reduction (80% CI: -2.34 – 4.04 %). Percentage reductions of deaths and hospitalizations for all possible distributions of self-tests are provided as Supporting Information.

This analysis shows that increasing the fraction of self-tests provided to the 19 − 64 age group could have averted more deaths and more hospitalizations. A potential explanation for this result is the fact that children in Greece have showed lower transmissibility compared to the middle-age group, which seems to have led the epidemic (3). The age-structure in the COVID-19 epidemic has been shown to be important in other country settings (4) suggesting that interventions at the middle-age group have more efficiency in reducing transmission compared to interventions at children.

#### Accuracy: Impact of the clinical accuracy of testing kits

Self-tests are naturally less accurate than PCR or antigen tests and the administration of the test by an individual instead of a medical professional could further diminish the self-tests credibility. In Figure 3, we present estimates on the percent changes in averted deaths and hospitalizations, relative to what is observed in the data, for different values of the sensitivity of self-tests. Our results indicate that higher quality tests would have contributed to averting more deaths and hospitalizations, albeit at significantly higher procurement costs and less availability (since higher quality self-tests were not available at the time of deployment).

**Fig. 3.**
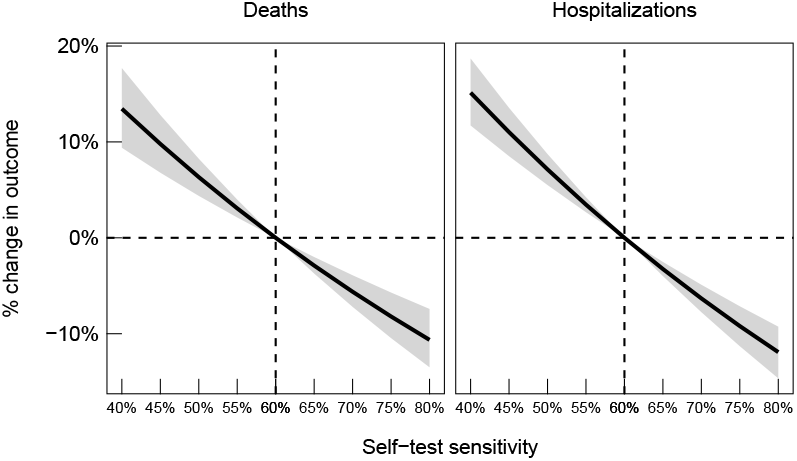
80% confidence intervals for percent changes in deaths (left panel) and hospitalizations (right panel), relative to what is observed in the data, as the sensitivity of self-tests is varied. Dashed line indicates the sensitivity reported by the manufacturers of the testing kits used in the Greek testing program.

**Fig. 4.**
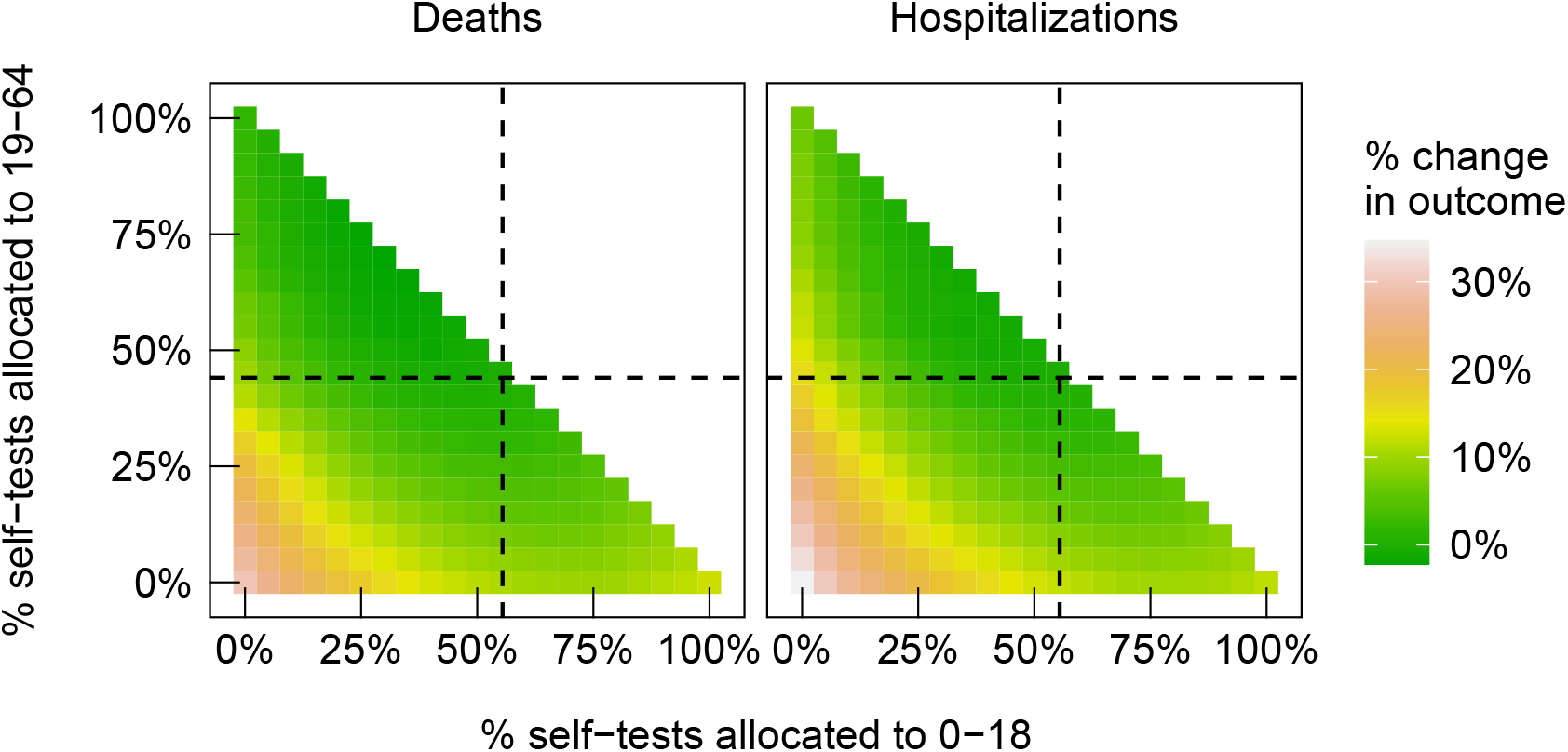
Heatmap of percent changes in deaths (left panel) and in hospitalizations (right panel), relative to what is observed in the data, as we vary the fractions of tests between different age groups: the 0–18 is allocated the percentage in the x-axis; the 19–64 age group is allocated the percentage in the y-axis; the 65+ group is allocated the remainder. Dashed lines indicate the fractions observed in the data.

**Fig. 5.**
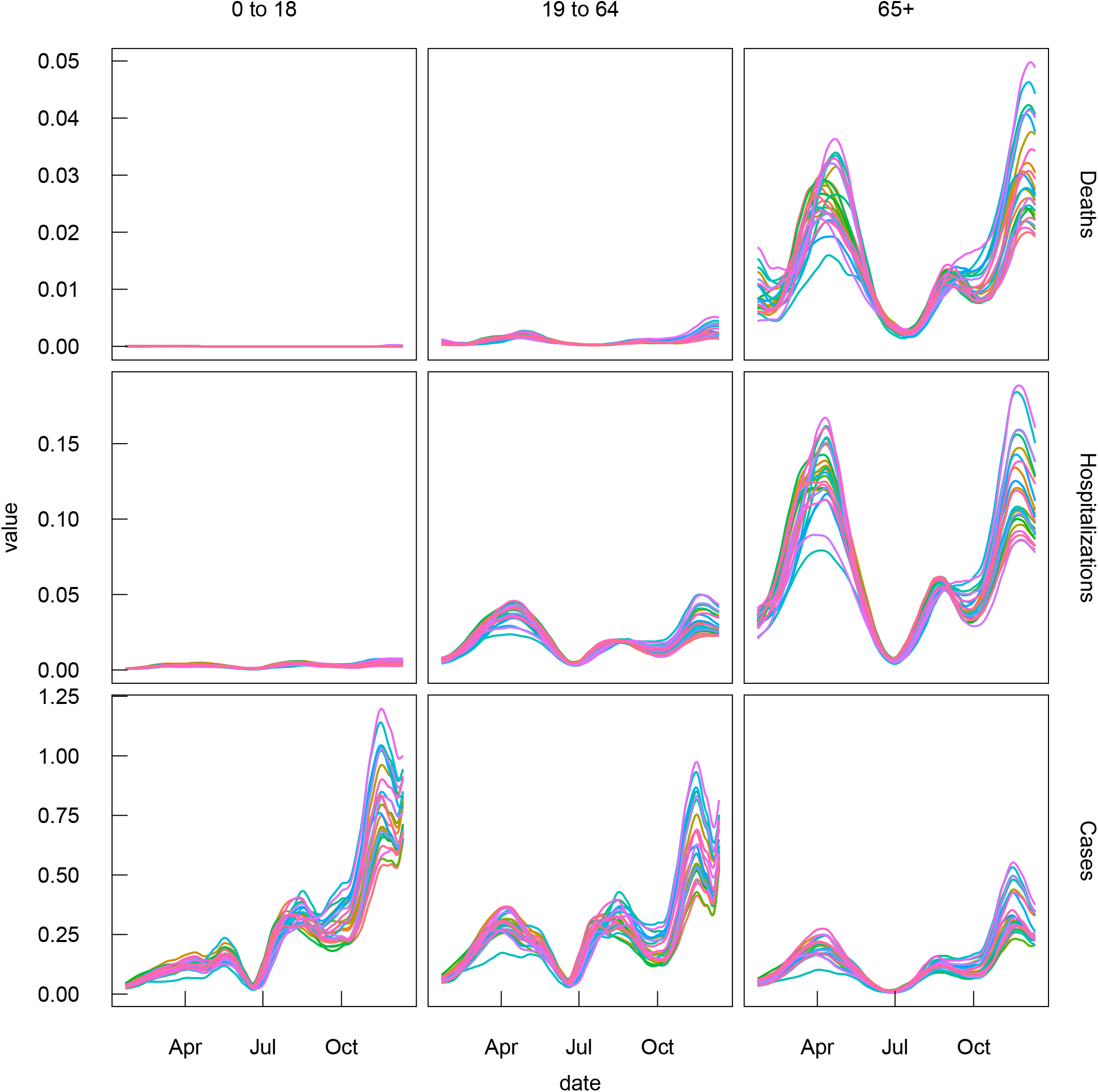
Deaths, hospitalizations, and cases per person split by age group. Each colored series corresponds to a time series from a single bootstrap sample.

**Fig. 6.**
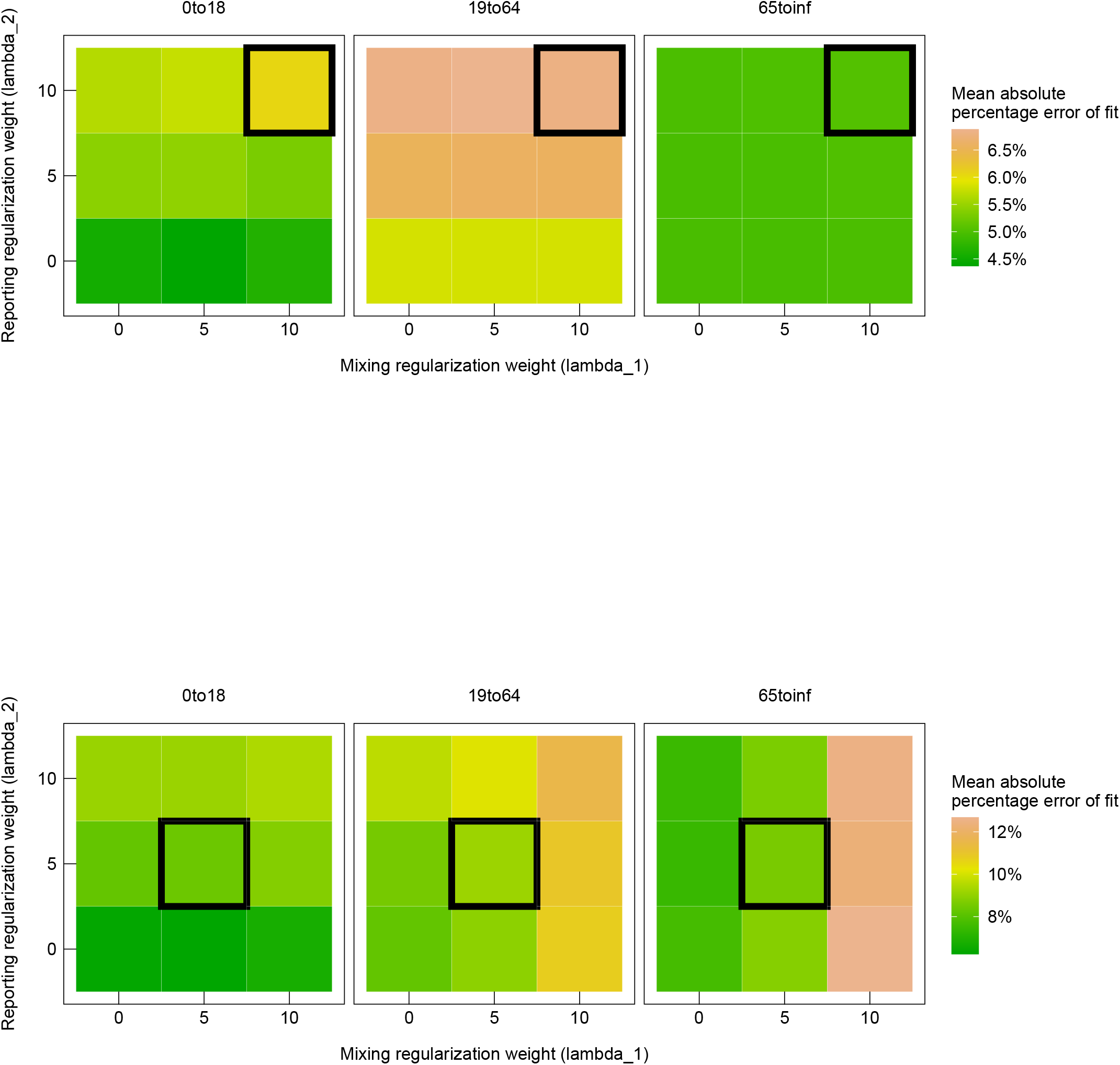
Median values of the mean absolute percentage achieved by the fitted model across all bootstrap datasets as the regularization weights vary, for both the first wave (top panel) and second wave (bottom panel). The selected weights were chosen to be as large as possible, whilst retaining a good fit to the data (low mean absolute percentage error).

## 2. Discussion

Our paper employs a compartmental model to investigate the effect of self-test based screening on the progression of an epidemic and the overall number of deaths and hospitalizations. Given that our analysis is based on observational data and not a (natural or designed) experiment, a number of assumptions are made that we document below. First, we assume that the dynamics of the disease follow the structure presented in Section 3. Such models are widely used in the mathematical epidemiology literature. Furthermore, when performing the sensitivity analysis we do not take into account other behavioural or policy changes caused by the perturbation that we evaluate, besides the potential test substitution effect we discussed in Section D. In other words, whenever a perturbation is evaluated we assume that everything else remains unaffected with the exception of the epidemic dynamics and potential test substitution. Moreover, false positive self-tests in susceptible individuals do not lead to them being isolated or hospitalized since in principle, they were followed up with a PCR or antigen test and resolved appropriately. Another assumption is that individuals die only after being hospitalized, implying that there are no deaths at home. Data fully supports this latter assumption, as there were almost no COVID-19-related deaths outside of a hospital in Greece. Finally, although individuals age, for simplicity we do not model any transitions between compartments of different age groups.

Despite the shortcomings of our approach, our model is rich enough to produce insights for policy makers and public health practitioners regarding the deployment and optimization of large scale screening programs. Concretely, we show that among the various NPIs large scale self-test screening is effective while less costly and associated with minimal societal and other implications. Furthermore, in order for such programs to be very effective scale is critical: testing a large percentage of the population daily (which implies testing individuals frequently) is a key component to success. Assuming that high population coverage is not feasible due to limited resources or prohibitive cost, targeting the most active subset of the population (age group 19 − 64) is more effective: our results indicate that this group is the main driver of the transmission. Finally, accuracy of the self tests is relevant and higher sensitivity contributes to non-trivial reduction of deaths and hospital admissions.

The pandemic of SARS-CoV-2 stressed the national health systems beyond their limits leading to severe disruptions of healthcare provision not only for COVID-19 patients, but also all other diseases. In the absence of therapeutic interventions, the goal of public health responses was initially to eliminate SARS-CoV-2. When elimination was realised as a non-realistic target and until vaccines were approved and deployed, the major goal was to decelerate transmission (5) of SARS-CoV-2 in levels where stress on the health systems would be manageable through Non Pharmaceutical Interven-tions (6).

Throughout the world a plethora of NPIs were implemented with different levels and combinations of social-distancing, face-cover wearing and screening (i.e., asymptomatic testing) programmes. Most NPIs with the exception of mass screening are associated with tremendous explicit (7) and implicit (8) costs while leading to a moderate impact on deaths and hospitalizations (2, 9). Slovakia (10) was the first country to test their entire population. This intervention led to a decrease in the reproduction number (and therefore the number of reported cases) shortly after the testing program (11) but this effect quickly disappeared because high-intensity testing was not sustainable (12).

Greece implemented a large-scale testing program that, in contrast to the Slovakia case, used self-tests which are not performed by medical professionals and return a result immediately albeit less accurate, which is considered secondary compared to fast turnaround and testing frequency (13). Self-testing emerged during COVID-19 pandemic firstly as a personal monitoring tool, but also as an important public health intervention to control transmission. Self-testing allows a person to perform the test in their own privacy and self-isolate within a very short-time period without waiting for results or having to move from their isolation. It also allows for testing to occur shortly before the the crucial event, thus minimizing the window between negative test and social contacts during which a person might be incubating the virus. The strength of large-scale self-testing intervention programs is that, as long as the distribution of the devices is secured, they can be easily scaled-up to include the whole population repeatedly and in a short-time with minimal cost. One criticism received during the deployment of self-testing devices described the possibility that positive results might not be reported back or even that self-test devices might not be used at all. However, this issue is shared with all screening interventions since all testing approaches need to take into account people’s consent and participation. Our analysis here, even with the most conservative estimates for prevention of hospitalisations and deaths, strongly supports that mass self-testing can provide a sustainable and efficient targeted public health intervention during pandemics.

The potential for future pandemics seems to be increasing as the global population grows and ages, but also as transportation is minimizing travel distances. Best practices during the COVID-19 pandemic should be documented and become part of strategic plans for pandemic preparedness. We have shown that mass screening through self-testing is one of these best practices that could provide an invaluable tool to decelerate the spread of a pandemic pathogen with minimal social and financial cost.

## 3. Materials and Methods

We provide details on the model we developed and the methods we employed in our analyses. We begin with a qualitative description of the compartments and transitions, and then elaborate on the model dynamics, i.e., on how we model the transitions using a mix of parameters and available historical data. Then, we discuss the methods we followed to fit the model, conduct sensitivity analyses, and evaluate the impact of the Greek mass screening program.

### A. Model Compartments and Transitions

Recall from the short description provided so far that we consider three sets of compartments based on age, 𝒢 = {‘0-18’,’19-64’,’65+’ }, and that we also consider two sets of compartments based on vaccination status, indexed by *v* ∈ {0, 1} to indicate vaccination.

Figure 1 illustrated, for a single age group *a* ∈ 𝒢, the model compartments and the possible transitions among them. As noted, there are two sets of compartments based on vaccination status. For the unvaccinated population, susceptible individuals (*S*_*a*0_) might transition to being vaccinated (*S*_*a*1_), or get infected. If infected, they transition to one of three compartments: asymptomatic and mild (*IAM*_*a*0_), symptomatic and mild (*ISM*_*a*0_), or symptomatic and severe (*ISS*_*a*0_). Populations in all three of these compartments can spread the disease to other populations that can get infected.

Infected individuals may be identified by taking a test, at which point those with mild disease move to an isolation compartment (*O*_*a*0_), whereas those with severe disease move to a hospitalization (*H*_*a*0_) compartment. Infected individuals with mild disease, however, might never be identified through testing, in which case they eventually recover into a compartment that continues to be subject to testing (*RT*_*a*0_). Infected individuals with severe disease are always eventually identified.

Populations in isolation with mild disease eventually recover, while hospitalized populations may recover or die; recovered individuals transition to (*R*_*a*0_), and dead to (*D*_*a*0_).

For the vaccinated population, transitions between compartments follow the same structure as with the unvaccinated, with the following exception: because vaccination could confer full immunity for a fraction of the vaccinated population, in the vaccinated compartments there is an additional transition that allows vaccinated individuals (*S*_*a*1_), after exposure, to directly recover into the compartment that continues to be subject to testing (*RT*_*a*1_).

### B. Model Dynamics and Data

Guided by the available data that is mostly derived from daily records, we consider discrete time steps that correspond to days. We index the time steps with *t* ∈ *{*0, 1, …, *T }*, where *T* is the model horizon. The dynamics of the model are described in terms of the number of individuals that transition between any two compartments in a single time step. To introduce some notation, let 𝒳 be the set of all compartments and 𝒳_*av*_ the set of all compartments for a given age group *a* and vaccination status *v*. For compartments *X, Y* ∈ 𝒳,we denote the number of individuals that transition from *X* to *Y* at time step *t* with Δ_X → Y_ (*t*). For each compartment *X* with population time at time step *t* equal to *X*(*t*), we then have the following dynamic update

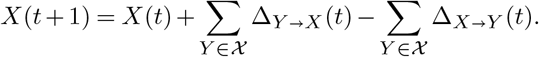

Certain transitions that we have historical data for are modeled explicitly using the data. Other transitions are modeled using parameters, which we then learn in the fitting procedure. To disambiguate, we adopt throughout the paper the following notation convention: quantities that are explicitly available using data are denoted with an overbar, whereas quantities that are to be learned by the fitting procedure are denoted with no overbar. For example, 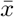 denotes a quantity that is explicitly provided in the data, whereas *x* is a quantity that needs to be learned.

We discuss the transitions in four groups: (i) Vaccination, (ii) Infection, (iii) Testing and Isolation, and (iv) Recovery, Hospitalization and Death.

#### (i) Vaccination

We refer to individuals for whom two doses are administered as vaccinated. We denote by 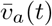 the recorded number of individuals in age group *a* who received their second dose on day *t*. The data are by the Greek National Public Health Organization (NPHO) records (14).

The available data does not specify whether individuals who got vaccinated at time *t* belonged in the Susceptible (*S*_*a*0_) or the Recovered compartment that is subject to testing (*RT*_*a*0_), which is why we assume a proportional split between the two, so that

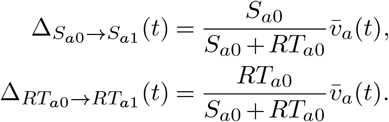

#### (ii) Infection

Infected individuals, by coming to close contact with others, might expose susceptible or vaccinated individuals to the virus and infect them. To model these infection dynamics, we seek first to quantify the total number of newly infected people, and, second, to quantify how these people transition into the different infection compartments in our model, depending on disease severity.

Let us focus on the transmissibility between two age groups, say *a, b* ∈ 𝒳, at time *t*. Consider an infected individual from age group *b*, and, to begin with, assume that all the population from age group *a* that they might come in contact with is unvaccinated and susceptible. Then, let *β*_*ab*_(*t*) be the number of people from age group *a* that the infected individual from age group *b* comes in close with and infects them. Note that these parameters, which we shall refer to as **mixing parameters**, capture, among others, the infectivity of the pathogen, the contagiousness of the infected individuals, and the contact patterns between different populations.

Eventually, at time step *t*, each infected individual from age group *b* need not infect as many as *β*_*ab*_(*t*) people from group *a*, because of two reasons. First, some of them might not be susceptible. Second, some of them might have developed immunity by being vaccinated.

Let us first focus on the unvaccinated people from group *a*. To calculate how many among them will eventually get infected due to an infected person from group *b*, we need to factor in the fraction of susceptible and unvaccinated among the population that the infected individual might come in contact with. The latter population, which we shall refer to as the **community population**, is drawn from all compartments, except from the Isolated, Hospitalized and Dead compartments. We denote the collection of aforementioned compartments with 𝒞_*a*_, i.e.,

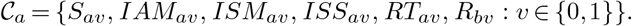

and denote the size of the associated community population with *C*_*a*_(*t*), with

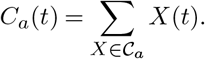

Using this notation then, each infected individual from age group *b* will infect *β*_*ab*_(*t*) × *S*_*a*0_(*t*)*/C*_*a*_(*t*).

To calculate how many unvaccinated people from group *a* will eventually get infected in total, we need to consider all infected populations from each age group. Let *I*_*b*_(*t*) be the number of infected individuals at time *t* from age group *b*, given by

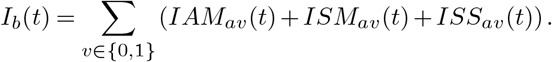

Letting *NI*_*av*_(*t*) be the total number of newly infected people from group *a* with vaccination status *v* at time *t*, we can now express *NI*_*a*0_(*t*) as

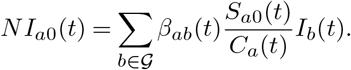

The breakthrough infection dynamics for the vaccinated people from group *a* are similar, with one difference: a fraction of vaccinated individuals who come in close contact with someone infected and would have otherwise been suject to a breakthrough infection, might not get infected at all due to vaccine-induced immunity. To model this, we introduce the **probability of vaccine immunity** 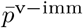. Therefore, a vaccinated individual who is exposed to the virus and would have been otherwise infected had they not been vaccinated, eventually does not get infected (cf. does get infected) with probability 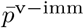 (cf. 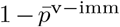. We assume this probability to remain the same across age groups. Note that this probability relates to the effectiveness of the vaccine.

(15) estimates vaccine effectiveness for the Omicron variant to be 80%. We adjust this value to 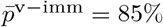 to take into account the higher vaccine effectiveness against non-Omicron variants that were prevalent in Greece throughout 2021 (accounting for vaccine types and variant proportions in the population (16)).

Using the probability of vaccine immunity, we can now express *NI*_*a*1_(*t*) as

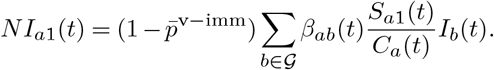

Newly infected people will develop symptoms and disease with varying severity. To model this, we introduce the following two parameters:

##### 1. Probability of Asymptomatic Disease

Let 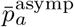 be the probability that an infected individual does not develop symptoms. We let this probability depend on age, and set:

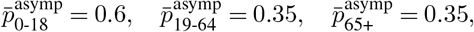

based on (17), who report these estimates for unvaccinated infected individuals. As there is no data to suggest that the proportion of asymptomatic infection is modified as a result of vaccination, we use the same probabilities for breakthrough infections as well.

##### 2. Probability of Severe Disease

Let 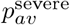 be the probability that an infected individual who is symptomatic, will also develop severe disease, as opposed to mild disease. Recall that those with severe disease eventually require hospitalization.

(18) provide estimates for the probabilities of infected and unvaccinated individuals who are hospitalized (1% for 0 to 18, 7% for 19 to 64, and 30% for 65+). These estimates do not quite correspond to 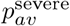 in our model, which is a conditional probability on symptomatic disease. Therefore, we let this as a free parameter to be learnt in the fitting process.

Putting all this together, we can express the transitions of susceptible population into the infection compartments as follows:

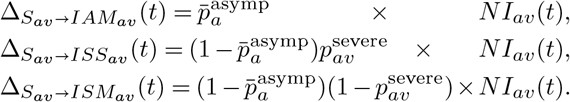

Finally, we note that the fraction of vaccinated susceptible individuals who come in close contact with someone infected but have developed vaccine-induced immunity, as we discussed previously, transition directly to the recovered compartment that continues to be subject to testing (*RT*_*a*1_). We can now express this transition as

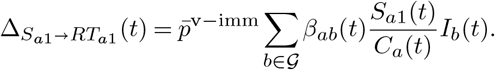

#### (iii) Testing and Isolation

Our model allows for two types of testing: regular tests and self tests. Both are conducted with the aim of isolating infected individuals, preventing them from spreading the disease or providing them with the appropriate care in the case of severe illness. We present each of the two types separately, since they affect the model dynamics differently.

**Regular tests** are in general more accurate than self-tests with clinical sensitivity and specificity given by 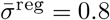 and 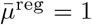, respectively. The available data from (19) provide the total number of regular tests administered in the country. We use Greek census data (20) as well as historical information to produce the number of regular tests performed daily to age group *a* and vaccination status *v*, denoted by 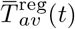, where we assume that tests are allocated proportional to the sizes of different age and vaccination groups.

The number of 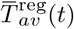 tests are split among different compartments. We next discuss how we model this split, letting 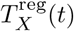 be the tests taken by population in compartment *X* ∈ 𝒳.

First, because newly hospitalized individuals were routinely tested upon admission, we set

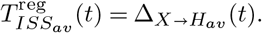

The remaining 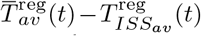 are assumed to be split as follows. Because regular tests were not required by the state, individuals in different compartments had different *propensities* to take them. We denote by *θ*_*a*_ and 1 − *θ*_*a*_ the testing propensity of asymptomatic and symptomatic individuals respectively and compute the *total propensity* of age group *a* and vaccination group *v*, denoted Θ_*av*_, as

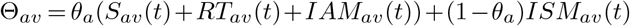

Then, we split the 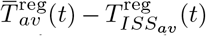 tests in proportion with the propensity and size of each compartment, i.e., we have that

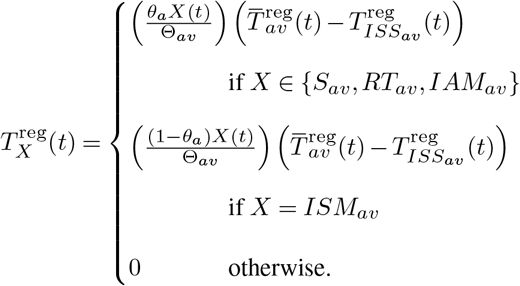

The total number of positive regular tests from compartment *X*, is therefore given by

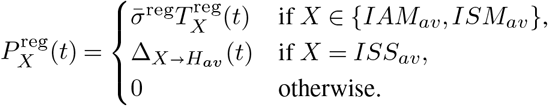

**Self tests** are in general less accurate than regular tests with clinical sensitivity and specificity given by 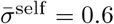 and 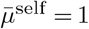, respectively. The available data from (19) provides the total number of self tests administered to a group (age group *a*, vaccination status *v*) for each week. We assume that these are uniformly distributed throughout the corresponding week and we denote by 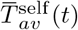 the number of self tests performed on day *t*.

Self tests are required by the state and therefore we can assume a uniform distribution amongst individuals within the eligible compartments. Therefore, the total number of self-tests administered to a compartment *X* can be calculated as

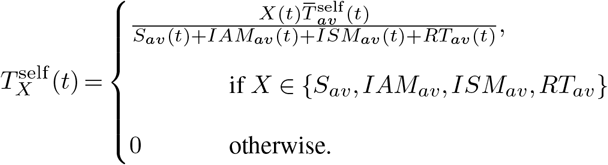

The total number of positive self-tests in each compartment is then given by

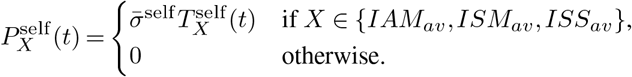

All individuals who are identified as positive and are not suffering from a severe infection move to the isolated compartment, i.e.,

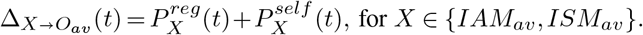

Those with a severe infection and identified as positive are hospitalized, as we discuss next.

#### (iv) Recovery, Hospitalization and Death

Infected individuals with mild disease spend an average of 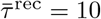 days (21) in the infected compartments (*IAM*_*av*_ or *ISM*_*av*_) before infectiousness subsides. Therefore, in the absence of a positive test, we have for each age group *a* and vaccination status *v*,

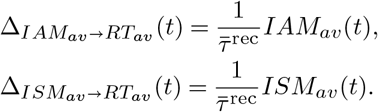

Similarly, we assume without loss that isolated individuals also spend an average of 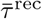 days before recovery, i.e.,

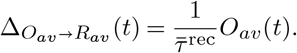

Infected individuals with severe disease, are hospitalized following 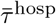 days after infection, i.e.,

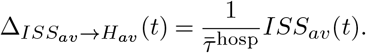

Hospitalised individuals are discharged either due to death or recovery. The average length of stay in the hospital, denoted by 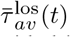, varies with age, vaccination, and month and is provided by the Greek NPHO (22). Among the discharged individuals, the fraction 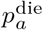 of those who die could be in principle estimated by the raw data. Unfortunately, the raw estimate varies substantially and therefore, we learn it through the fitting process. Combining the above, we have the following transitions

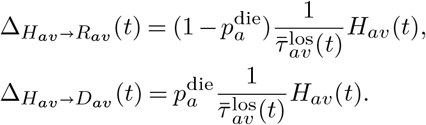

### C. Model Fitting

We fit the model against the following key outcomes, for which we have historical data: daily recorded hospitalizations, deaths, total cases, and cases reported through the self-testing program. We then find the parameter values that minimize the sum of squared log errors between our model’s predictions for these outcomes and the data. We elaborate on the key outcomes and data next, and detail the minimization procedure in the Supporting Information.

#### Hospitalizations and Deaths

The number of daily hospitalizations and deaths by age group are available from data recorded by the NPHO of Greece (23). The corresponding model predictions are calculated by

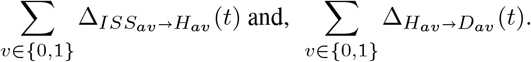

#### Total Cases

The number of daily total new cases by age group are available from data recorded by the Greek NPHO (24). The corresponding model predictions are given by

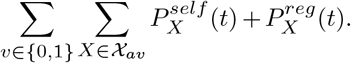

#### Cases reported through the self-testing program

The number of daily new cases by age group from the self-testing program are available from data recorded by (24). Notably, these cases were predominantly recorded following a positive self test. In addition, it is suspected that some of these cases were erroneously recorded through the program following a positive regular test, in the absence of a positive self test. To account for this possibility, let *γ*_*a*_(*t*) be the fraction of regular positive tests for age group *a* at time *t* that were reported through the self-test program. The model’s prediction of the number of cases reported through the self-testing program is equal to

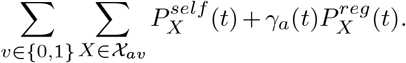

Note that *γ*_*a*_(*t*) is assumed to vary with age and over time, to account for different types of reporting errors as the system processes varying amounts of self tests in the population. The data we fit the model to and expressions for the corresponding model predictions are summarized below:

- Hospitalizations: 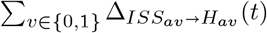
- Deaths: 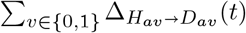
- Total Cases: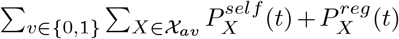
- Cases through Self-Tests Program: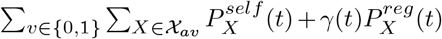

### D. Sensitivity Analyses

For each sensitivity analysis, we fit the model to the data and then simulate it for varying values of the parameter of interest, keeping other parameters constant and equal to their fitted values. We perform these simulations within a bootstrapping process, so as to obtain not just point estimates, but confidence intervals as well. We discuss how we performed the simulations below, and detail the bootstrapping process in the Supporting Information.

To estimate the impact of the number of self tests deployed, in the simulations we vary the number of self tests taken by scaling them uniformly across time and different population groups. That is, at time *t*, we consider the number of self tests taken by population in age group *a* and vaccination status *v* to be 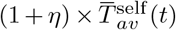, where *η* is a scaling parameter. For *η* = 0, we recover the baseline scenario that was implemented in reality; for *η <* 0 (cf. *η >* 0), we consider scenarios in which the scale of the program is decreased (cf. increased). A self-testing program of a different scale would have likely induced a testing behavior of the population different than the one observed in the baseline scenario. If self tests were reduced, for example, some individuals who self tested in reality and could no longer test under this reduced-scale scenario, might seek to take a regular test instead, particularly if they were symptomatic. Ignoring this potential behavioral change could overstate the effect of self tests, especially when self tests are reduced, i.e., for *η <* 0. To alleviate this concern, we make the following modification to the simulation. First, we compute, under the baseline scenario, for individuals within a given community compartment *X* ∈ 𝒞_*a*_, the fraction of them who seeked a regular test among those who did not take a self test, i.e.,

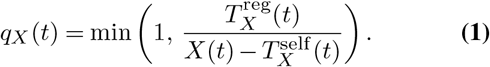

The fraction *q*_*X*_ (*t*) then corresponds to the propensity of the population in *X* to take a regular test when they are unable to take a self test. We assume that this propensity would remain the same, regardless of the self-testing program’s scale. Let 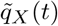 be the propensities that are produced after fitting the model to the data. Then, in the simulations for our sensitivity analysis, we modify the number of regular tests taken to be

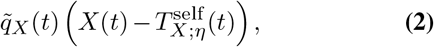

where 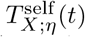 is the number of self-tests in compartment *X*, after the total number of self-tests are scaled by (1 + *η*).

To estimate the impact of the self test allocation mix among different age groups, we consider varying the fractions of total tests allocated to each group—subject to the fractions summing of to one, so that to total number of tests is held constant. For each set of fractions we consider, we scale the number of tests taken uniformly within each age group accordingly.

Finally, for the sensitivity analysis on the test’s clinical sensitivity, we simply use the fitted parameters to simulate the outputs of the model for various test sensitivity values 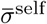.

### E. Overall Impact of the Program

To evaluate the overall impact of the program on deaths and hospitalizations, we consider two methodologies, both of which rely on our sensitivity analysis on the number of self tests taken that we described above. We refer to them as direct and indirect methods:

1. Direct method: we conduct the analysis by simply setting *η* = − 1, which corresponds to the scenario of no self tests conducted, and obtain estimates for deaths and hospitalizations.
2. Indirect method: we first use the analysis to estimate local derivatives at the baseline scenario for deaths and hospitalizations, which we denote with ∂*D* and ∂*H*. In particular, we run the analysis for *η* = − 0.01 and *η* = ± 0.01, i.e., we consider ±1% perturbations of the program’s scale, and then use finite differences to estimates ∂*D* and ∂*H*. Given that the total number of deaths and hospitalization are concave in the number of self tests conducted, i.e., additional self tests exhibit diminishing returns, conservative estimates of the total number of deaths and hospitalization are provided by

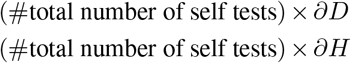

Finally, to evaluate the overall impact of the program on transmissibility, we calculate the effective reproduction number *R*_*t*_ for each time step for the fitted model by using the methodology in (1). Then, we follow the sensitivity analysis on the number of self tests taken by setting *η* = − 1, which corresponds to the scenario of no self tests conducted, and obtain estimates for *R*_*t*_ in a similar fashion.

## Data Availability

All data produced in the present study are available upon reasonable request to the authors

## 4. Appendix

### Additional Program Implementation Details

In March 2021, when another wave of the COVID-19 pandemic was putting high pressure to the Greek National Health System, the authorities decided to launch mandatory weekly testing starting with students in all schools. The self-testing kits were distributed through pharmacies across the country, on a weekly basis, with the use of social security numbers. Control was carried out either with the display of a solemn declaration stating that the individual was tested negative, or through an online form available at self-testing.gov.gr. For cases of positive self-test results, an amendment of legislation imposed the implementation of a second test—either PCR or antigen-test––by a health-care professional, to confirm the first test’s result and update the National Registry for Covid-19 Patients. The authorities also proceeded occasionally with the distribution of free self-testing kits to the entire population, particularly before or after public holidays.

The distribution of self-testing kits and the implementation of mandatory checks began on 07-04-2021 on the re-opening of High Schools, Vocational High Schools and Junior High Schools as well as Junior High Schools with Senior Classes, with the provision of 2 self-testing kits per week, for all students as well as the schools’ teaching and other staff.

On 19-04-2021, the government passed a law imposing mandatory self-testing also for private sector employees and civil servants working at their workplace. Self-testing kits were distributed to employees throughout the summer, while distribution to teaching staff and students stopped during the summer holidays.

In September 2021, the state decided to impose a once-a-week mandatory self-testing at own cost for private sector employees and civil servants who were not fully vaccinated or who have not had COVID-19. With the beginning of the new school year, weekly self-testing was mandatory only for students aged 4-18 who were not vaccinated or have not had COVID-19. It should be noted that when students were considered as “close contacts” of a COVID-19 case, the Education Division would provide additional self-tests to ensure daily testing.

Finally, private sector employees and civil servants who were not vaccinated or have not had COVID-19 were obliged to carry out two weekly tests from 05-11-2021 onwards, at their own cost, considering the role of seasonality in the spread of COVID-19 pandemic.

During the aforementioned period, a total of 97,000,000 self-testing kits were purchased and distributed first to the country’s 100 pharmaceutical warehouses and thereafter to local pharmacies.

### Minimization

To fit the model, we find the parameter values that minimize the sum of squared log errors between our model’s predictions for the outcomes we discussed in the Model Fitting section and the data.

To introduce some notation, consider some age group *a* and some time period *t*. Let 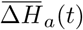 and 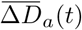 be the corresponding number of new hospitalizations and deaths, respectively; let 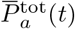 be the corresponding total number of cases recorded; and let 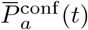 be the corresponding number of cases reported through the self-testing program. The data and corresponding predictions are summarized in Table 2.

**Table 2.**
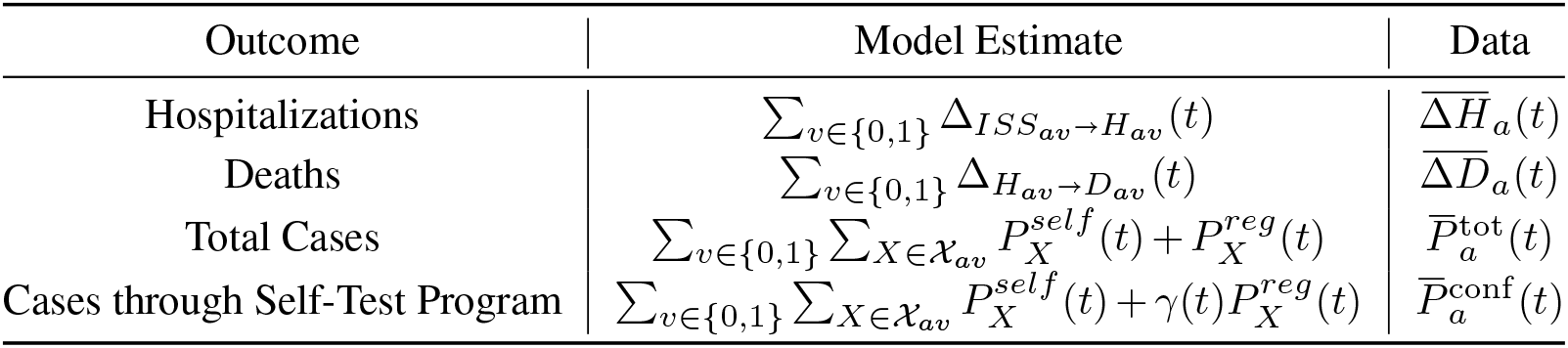
Model estimates and observed data used for model fitting.

| Outcome | Model Estimate | Data |
| --- | --- | --- |
| Hospitalizations | $\sum_{v \in \{0,1\}} \Delta_{ISS_{av} \rightarrow H_{av}}(t)$ | $\overline{\Delta H}_a(t)$ |
| Deaths | $\sum_{v \in \{0,1\}} \Delta_{H_{av} \rightarrow D_{av}}(t)$ | $\overline{\Delta D}_a(t)$ |
| Total Cases | $\sum_{v \in \{0,1\}} \sum_{X \in \mathcal{X}_{av}} P_X^{self}(t) + P_X^{reg}(t)$ | $\overline{P}_a^{tot}(t)$ |
| Cases through Self-Test Program | $\sum_{v \in \{0,1\}} \sum_{X \in \mathcal{X}_{av}} P_X^{self}(t) + \gamma(t) P_X^{reg}(t)$ | $\overline{P}_a^{conf}(t)$ |

We use a non-convex optimization algorithm to minimize the sum of squared log errors between our model’s prediction and the target data. In order to avoid overfitting we add a regularization term in the loss function.

#### A. Parameters and Initialization

The parameters that we fit are summarized in Table 3. To facilitate the fitting process, we constrain certain model parameters as we discuss next, and summarized in Table 3.

**Table 3.**
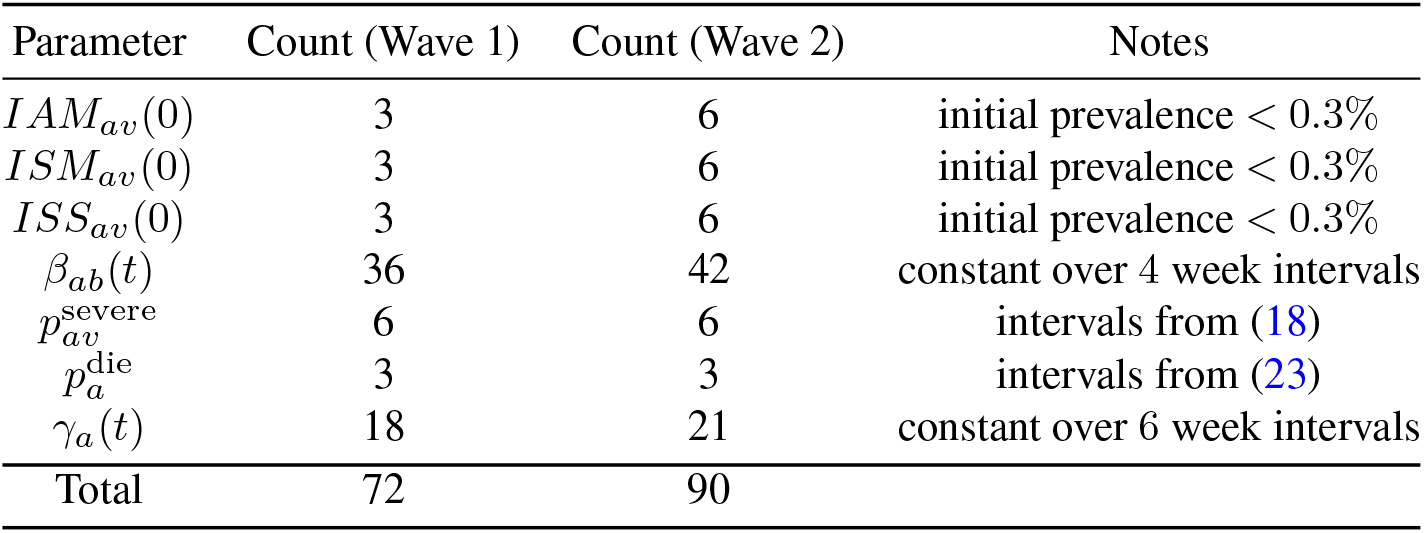
Number of parameters learned by the model in each of the two waves. We use a sparsity term in the objective to prevent overfitting.

The probabilities of severe disease 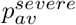, though learned from the fitting process defined below, are constrained to lie within the following intervals

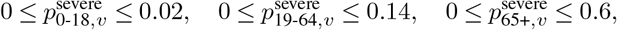

which have widths twice the size of the estimates in (18). Similarly, we learn 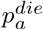 and constrain them to lie in intervals [0, 2*q*_*a*_] where *q*_*a*_ represents the total deaths divided by the total hospitalizations in age group *a* in a given wave, obtained from (23). The mixing parameters *β*_*ab*_(*t*) and the reporting parameters *γ*_*a*_(*t*) are assumed to be constant in 4 and 6 week intervals respectively. These limits were empirically established, with the goal of enabling tractability and avoiding overfitting.

We use data between January 21, 2021 until December 15, 2021. During the said period, Greece experienced two epidemic waves, one lasting from January 21, 2021 until June 20, 2021, and the other from June 21, 2021 until December 15, 2021. The self-testing program was introduced in the middle of the first of these waves and continued through the second wave. We fit the model for these two waves. For the fist wave, the sizes of all compartments are initialized at 0 with the following exceptions. The number of initially hospitalized patients are directly estimated from the raw data. The size of the susceptible compartments and infected compartments are allowed to be nonzero (and are learned through the fitting process) subject to the following constraints: (i) the prevalence is assumed to be less than 0.3%, where this upper bound is obtained by (25), (ii) the sum of all compartments sums to the population data per age group provided in (20).

For the second wave, the sizes of all compartments are initialized at their levels learned from the first wave fitting process, with the exception of the infected compartments that are learned through the second wave’s fitting process. Similar to the first wave, we impose the constraint that the sum of all compartments sums to the population data per age group provided in (20). Note that fitting the sizes of the infected compartments for the second wave is necessary, because the first-wave fit for these quantities at the end of the first wave might not be credible owing to end-of-horizon effects.

#### B. Loss Function

We consider a loss function that is the sum of squared log-errors, across time steps, age groups, and waves for all outcomes presented in Table 2. There are more than 100 parameters to fit and hence the loss function is complex with no analytical gradients, making the underlying optimization problem a difficult learning task with the potential risk of overfitting. In order to obtain sparse solutions, we proceed by using regularization on the time-varying parameters (*β*_*ab*_(*t*) and *γ*_*a*_(*t*)) and a two-stage block-minimization technique to ease the optimization burden.

The regularization is a standard penalty on the absolute differences in successive values of the time-varying parameters. In particular, we introduce two regularization parameters, *λ*_1_ and *λ*_2_ and we add the following component to the loss function

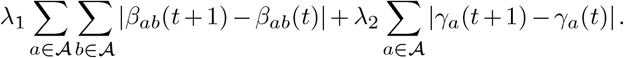

The block-minimization approach works in two steps:

Step 1: We numerically minimize the regularized loss function with the cross-group mixing parameters fixed to zero (*β*_*ab*_(*t*) = 0 for *a* ≠ *b*), and obtain an initial estimate on the unknown parameters.

Step 2: We numerically minimize the loss function by varying all mixing parameters, *β*_*ab*_(*t*), keeping the rest fixed to their values from Step 1.

To complete both fitting steps we use the Levenberg-Marquardt algorithm implemented in (26), and add random restarts. The regularization parameters are fixed in the optimization and chosen by searching through a grid. All computational experiments are run on the SuperCloud infrastructure (27).

#### Bootstrapping for Confidence Intervals

To derive confidence intervals for our sensitivity analyses, we follow an approach that is similar to traditional bootstrapping. It relies on the data being split according to the 74 regional units of Greece, and uses this split to construct “virtual” populations that were obtained by randomly sampling regional units (with replacement) until the total population of the collection was at least 2 million. We observed that the time series within individual regional units with small populations suffered from high variance, and therefore the 2 million resident threshold was selected so that the time series of the virtual populations were stable. Figure 5 shows the profiles of the 25 bootstrap samples.

In our analysis we set *N* = 25 for tractability, and choose to report 80% confidence intervals. Confidence intervals for the quantities of interest are derived by completing the fitting procedure and analysis on each of these datasets.

#### Footnotes

1 https://www.finddx.org

## Bibliography

1. Francisco Arroyo-Marioli, Francisco Bullano, Simas Kucinskas, and Carlos Rondón-Moreno. Tracking r of covid-19: A new real-time estimation using the kalman filter. PloS one, 16(1): e0244474, 2021.

2. S. Flaxman, S. Mishra, A. Gandy, H. J. T. Unwin, T. A. Mellan, H. Coupland, C. Whittaker, H. Zhu, T. Berah, J. W. Eaton, M. Monod, “Imperial College COVID-19 Response Team”, Ghani A. C.and Donnelly C. A.and Riley S.and Vollmer M. A. C.and Ferguson N. M.and Okell L. C., and S. Bhatt. Estimating the effects of non-pharmaceutical interventions on covid-19 in europe. Nature, 584:56–71, 2020.

3. Spyros Sapounas, Angeliki Bistaraki, Edison Jahaj, Anastasia Kotanidou, Pagona Lagiou, and Gkikas Magiorkinis. Cold-season epidemic dynamics of covid-19 in two major metropolitan areas in greece: Hypotheses and implications for public health interventions. Frontiers in Medicine, 9, 2022. ISSN 2296-858X. doi: 10.3389/fmed.2022.861185.

4. Mélodie Monod, Alexandra Blenkinsop, Xiaoyue Xi, Daniel Hebert, Sivan Bershan, Simon Tietze, Marc Baguelin, Valerie C. Bradley, Yu Chen, Helen Coupland, Sarah Filippi, Jonathan Ish-Horowicz, Martin McManus, Thomas Mellan, Axel Gandy, Michael Hutchinson, H. Juliette T. Unwin, Sabine L. van Elsland, Michaela A. C. Vollmer, Sebastian Weber, Harrison Zhu, Anne Bezancon, Neil M. Ferguson, Swapnil Mishra, Seth Flaxman, Samir Bhatt, Oliver Ratmann, and null null. Age groups that sustain resurging covid-19 epidemics in the united states. Science, 371(6536):eabe8372, 2021. doi: 10.1126/science.abe8372.

5. Berber Snoeijer, Mariska Burger, Shaoxiong Sun, Richard Dobson, and Amos Folarin. Measuring the effect of non-pharmaceutical interventions (npis) on mobility during the covid-19 pandemic using global mobility data. npj Digital Medicine, 4, 12 2021. doi: 10.1038/s41746-021-00451-2.

6. Nikolaos Askitas, Konstantinos Tatsiramos, and Bertrand Verheyden. Estimating worldwide effects of non-pharmaceutical interventions on covid-19 incidence and population mobility patterns using a multiple-event study. Scientific Reports, 11:1972, 01 2021. doi: 10.1038/s41598-021-81442-x.

7. Antoine Mandel and Vipin Veetil. The economic cost of covid lockdowns: An out-of-equilibrium analysis. Economics of Disasters and Climate Change, 4, 10 2020. doi: 10.1007/s41885-020-00066-z.

8. Anton Pak, Oyelola Adegboye, and Emma Mcbryde. Are we better-off? the benefits and costs of australian covid-19 lockdown. Frontiers in Public Health, 9, 12 2021. doi: 10.3389/fpubh.2021.798478.

9. Sebastian Mader and Tobias Rüttenauer. The effects of non-pharmaceutical interventions on covid-19 mortality: A generalized synthetic control approach across 169 countries. Fron-tiers in Public Health, 10:820642, 04 2022. doi: 10.3389/fpubh.2022.820642.

10. Ed Holt. Slovakia to test all adults for sars-cov-2. Lancet, 396:1386–1387, 2020. doi: 10.1016/S0140-6736(20)32261-3.

11. Jaroslav Frnda and Marek Ďurica. On pilot massive covid-19 testing by antigen tests in europe. case study: Slovakia. Infectious Disease Reports, 13:45–57, 01 2021. doi: 10.3390/idr13010007.

12. Martin Pavelka, Kevin Van-Zandvoort, Sam Abbott, Katharine Sherratt, Marek Majdan, null null, null null, Pavol Jarčuška, Marek Krajčí, Stefan Flasche, and Sebastian Funk. The im-pact of population-wide rapid antigen testing on sars-cov-2 prevalence in slovakia. Science, 372(6542):635–641, 2021. doi: 10.1126/science.abf9648.

13. Daniel B. Larremore, Bryan Wilder, Evan Lester, Soraya Shehata, James M. Burke, James A. Hay, Milind Tambe, Michael J. Mina, and Roy Parker. Test sensitivity is secondary to frequency and turnaround time for covid-19 screening. Science Advances, 7(1): eabd5393, 2021. doi: 10.1126/sciadv.abd5393.

14. NPHO of Greece. COVID-19 Vaccinations in Greece [Unpublished raw data], 2021.

15. Tommy Nyberg, Neil M Ferguson, Sophie G Nash, Harriet H Webster, Seth Flaxman, Nick Andrews, Wes Hinsley, Jamie Lopez Bernal, Meaghan Kall, Samir Bhatt, et al. Comparative analysis of the risks of hospitalisation and death associated with sars-cov-2 omicron (b. 1.1. 529) and delta (b. 1.617. 2) variants in england: a cohort study. The Lancet, 399(10332): 1303–1312, 2022.

16. Toon Braeye, Lucy Catteau, Ruben Brondeel, Joris A.F. van Loenhout, Kristiaan Proesmans, Laura Cornelissen, Herman Van Oyen, Veerle Stouten, Pierre Hubin, Matthieu Billuart, Achille Djiena, Romain Mahieu, Naima Hammami, Dieter Van Cauteren, and Chloé Wyndham-Thomas. Vaccine effectiveness against onward transmission of sars-cov2-infection by variant of concern and time since vaccination, belgian contact tracing, 2021. Vaccine, 40(22):3027–3037, 2022. ISSN 0264-410X. doi: https://doi.org/10.1016/j.vaccine.2022.04.025.

17. Qiuyue Ma, Jue Liu, Qiao Liu, Liangyu Kang, Runqing Liu, Wenzhan Jing, Yu Wu, and Min Liu. Global percentage of asymptomatic sars-cov-2 infections among the tested population and individuals with confirmed covid-19 diagnosis: a systematic review and meta-analysis. JAMA network open, 4(12):e2137257–e2137257, 2021.

18. Tjede Funk, Francesco Innocenti, Joana Gomes Dias, Lina Nerlander, Tanya Melillo, Charmaine Gauci, Jackie M Melillo, Patrik Lenz, Helena Sebestova, Pavel Slezak, et al. Agespecific associations between underlying health conditions and hospitalisation, death and in-hospital death among confirmed covid-19 cases: a multi-country study based on surveillance data, june to december 2020. Eurosurveillance, 27(35):2100883, 2022.

19. NPHO of Greece. Self-Testing Figures and Cases Confirmed Positive [Unpublished raw data], 2021.

20. Hellenic Statistical Authority. 2011 Population-Housing Census. https://www.statistics.gr/el/statistics/-/publication/SAM03/2011, 2011.

21. Andrew William Byrne, David McEvoy, Aine B Collins, Kevin Hunt, Miriam Casey, Ann Barber, Francis Butler, John Griffin, Elizabeth A Lane, Conor McAloon, et al. Inferred duration of infectious period of sars-cov-2: rapid scoping review and analysis of available evidence for asymptomatic and symptomatic covid-19 cases. BMJ open, 10(8):e039856, 2020.

22. NPHO of Greece. Length of Hospital Stays for COVID-19 Patients in Greece [Unpublished raw data], 2021.

23. NPHO of Greece. Hospitalizations and Deaths for COVID-19 Patients in Greece [Unpub-lished raw data], 2021.

24. NPHO of Greece. Daily Covid-19 Cases. https://eody.gov.gr/category/covid-19/, 2021.

25. Gupta V. et al. Bastani H., Drakopoulos K. Efficient and targeted COVID-19 border testing via reinforcement learning. Nature, 599:108–113, 2021.

26. Matt Newville, Renee Otten, Andrew Nelson, Antonino Ingargiola, Till Stensitzki, Dan Allan, Austin Fox, Faustin Carter Michal, Ray Osborn, Dima Pustakhod lneuhaus, Sebastian Weigand Glenn, Christoph Deil Mark, Allan L. R. Hansen, Gustavo Pasquevich, Leon Foks, Nicholas Zobrist, Oliver Frost, Alexandre Beelen, Stuermer, azelcer, Andrew Hannum, Anthony Polloreno, Jens Hedegaard Nielsen, Shane Caldwell, Anthony Almarza, and Arun Persaud. lmfit/lmfit-py: 1.0.3, October 2021.

27. Albert Reuther, Jeremy Kepner, Chansup Byun, Siddharth Samsi, William Arcand, David Bestor, Bill Bergeron, Vijay Gadepally, Michael Houle, Matthew Hubbell, Michael Jones, Anna Klein, Lauren Milechin, Julia Mullen, Andrew Prout, Antonio Rosa, Charles Yee, and Peter Michaleas. Interactive supercomputing on 40,000 cores for machine learning and data analysis. In 2018 IEEE High Performance extreme Computing Conference (HPEC), pages 1–6. IEEE, 2018.

